# Proteome-wide association studies using summary pQTL data of three tissues identified 30 risk genes of Alzheimer’s disease dementia

**DOI:** 10.1101/2024.03.28.24305044

**Authors:** Tingyang Hu, Qiang Liu, Qile Dai, Randy L. Parrish, Aron S. Buchman, Shinya Tasaki, Nicholas T. Seyfried, Yanling Wang, David A. Bennett, Philip L. De Jager, Michael P. Epstein, Jingjing Yang

**Author notes:** Co-first authors (Tingyang Hu, Qiang Liu). Correspondence to: Jingjing Yang, Center for Computational and Quantitative Genetics, Department of Human Genetics, Emory University School of Medicine, Atlanta, GA, 30322, USA.

## Abstract

**Background:** Proteome-wide association study (PWAS) integrating proteomic data with genome-wide association study (GWAS) summary data is a powerful tool for studying Alzheimer’s disease (AD) dementia. Existing PWAS analyses of AD often rely on the availability of individual-level proteomic and genetic data of a reference panel. Leveraging summary protein quantitative trait loci (pQTL) reference data of multiple AD-relevant tissues is expected to improve PWAS findings of AD dementia.

**Methods:** We conducted PWAS of AD dementia by integrating publicly available summary pQTL data of brain, cerebrospinal fluid (CSF), and plasma tissues, with the latest GWAS summary data of AD dementia. For each target protein per tissue, we employed our recently published OTTERS tool to obtain omnibus PWAS p-value, to test whether the genetically regulated protein abundance in the corresponding tissue is associated with AD dementia. Protein-protein interactions and enriched pathways of identified significant PWAS risk genes were analyzed by STRING. The potential causal effects of these PWAS risk genes were assessed by probabilistic Mendelian randomization analyses.

**Results:** We identified 30 unique significant PWAS risk genes for AD dementia, including 11 for brain, 9 for CSF, and 16 for plasma tissues. Four of these were shared by at least two tissues, and gene *MAPK3* was found in all three tissues. We found that 11 of these PWAS risk genes were associated with AD or AD pathological hall marks as shown in GWAS Catalog; 18 of these were detected by transcriptome-wide association studies (TWAS); and 25 of these, including 8 out of 9 novel genes, were interconnected within a protein-protein interaction network involving the well-known AD risk gene *APOE*. Especially, these PWAS risk genes were enriched in immune response, glial cell proliferation, and high-density lipoprotein particle clearance pathways. Mediated causal effects were validated for 13 PWAS risk genes (43.3%).

**Conclusions:** Our findings provide novel insights into the genetic mechanisms of AD dementia in brain, CSF, and plasma tissues, and targets for developing therapeutic interventions. We also demonstrated the effectiveness of integrating summary pQTL and GWAS data for mapping risk genes of complex human diseases.

## Background

Large-scale genome-wide association studies (GWAS) have successfully identified dozens of genetic risk loci related to Alzheimer’s disease (AD) dementia^1–3^. However, the underlying molecular mechanisms of these GWAS risk genes of AD are still largely unknown. To gain biological insights into how associated risk genes might contribute to AD dementia, researchers have performed proteome-wide association studies (PWAS) that integrates reference proteomic data from an AD-related tissue with GWAS summary data of AD dementia to identify risk genes whose effects are mediated via genetically regulated protein abundance^4,5^.

PWAS typically employs a two-stage framework: Stage I uses the genetic and proteomic data of the same reference cohort to train a protein abundance prediction model for each target protein, taking the protein abundance quantitative trait as the response variable and the cis-genetic variants proximal to the protein-coding gene as predictors. The estimated genetic coefficients from Stage I can be viewed as effect sizes of “protein quantitative trait loci (pQTL)” in a broad sense, as most genetic variants with non-zero effect sizes will not be statistically significant pQTL. Stage II proceeds by using the estimated pQTL effect sizes as variant weights to predict genetically regulated protein abundance in a GWAS cohort, and subsequently conducts a gene-based association test (of the corresponding protein-coding gene) relating the predicted abundance of the target protein to phenotype.

Existing analytic tools derived for the analogous transcriptome-wide association studies (TWAS) have been used for PWAS. Existing tools such as TIGAR^6^, PrediXcan^7^, and FUSION^8^, utilize different statistical methods in Stage I to estimate the pQTL weights, requiring individual-level genetic and proteomic data of the reference cohort. For example, by PWAS analyses of AD dementia with the individual-level reference proteomic data of dorsolateral prefrontal cortex (DLPFC) tissue and whole genome sequencing (WGS) genotype data from samples in the Religious Orders Study and Rush Memory and Aging Project (ROS/MAP)^9^, Wingo et al.^4^ detected 11 risk genes by using the FUSION^8^ tool alone and Hu et al.^10^ identified 43 risk genes by aggregating results obtained from three tools of TIGAR^6^, PrediXcan^7^, and FUSION^8^.

In this work, we utilized our newly developed OTTERS^11^ tool to extend the PWAS analyses of AD dementia, by leveraging the summary pQTL data that was recently released and publicly available. This data includes not only brain (parietal lobe cortex, n=380) but also cerebrospinal fluid (CSF, n=835) and plasma (n=529) tissues^12^. Recent studies have shown that amyloid beta (A*β*)1-42/A*β*1-40 and phosphorylated tau/A*β*1-42 ratios in CSF^13,14^ and plasma^15,16^ could be used as biomarkers for early diagnosis of AD. Both CSF and plasma tissues are AD-relevant and important for studying genetic mechanisms of AD dementia. Thus, conducting PWAS with the recent GWAS summary data of AD dementia (n=∼789K)^3^ in all three tissues (brain, CSF, and plasma) is expected to identify additional risk genes of AD whose genetic effects are potentially mediated through the genetically regulated protein abundances.

## Methods

### OTTERS framework

In a two-stage PWAS framework with individual-level genetic and proteomic data from a reference panel, Stage I involves fitting a multiple linear regression model (*Equation 1*) with protein abundance (**E**_*p*_) of a protein *p* as the outcome, genotype data (**X**) of cis-genetic variants of the corresponding protein-coding gene (i.e., genetic variants located within the ±1Mb region around gene transcription start/termination sites) as predictors, and ***w*** denoting the pQTL weights to be estimated:

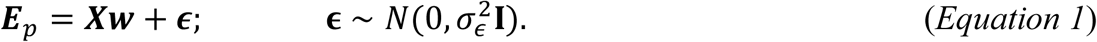

Potential confounding covariates are assumed to be adjusted from the original protein abundance measures, resulting in the residuals ***E***_*p*_. Both ***E***_*p*_ and columns of **X** are standardized with mean 0 and variance 1.

When individual-level genetic and proteomic data from a reference panel are not available, OTTERS^11^ can still estimate **w** in *Equation 1* by using only the summary pQTL reference data that are generated based on the following single-variant linear regression models with standardized genotype vectors **x**_*j*_ for genetic variants *j* = 1, …, *m*:

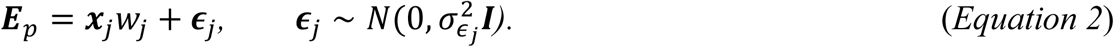

Summary pQTL reference data include the marginal least squared effect estimates (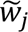, *j* = 1, …, *m*), sample size, and linkage disequilibrium coefficients in the reference pQTL cohort (which can also be approximated by using an external reference panel with the same ancestry). OTTERS employs five representative PRS models, including the P-value Thresholding with linkage disequilibrium (LD) clumping (*P+T*)^17^ with p-value thresholds of 0.05 (*P+T_0.05*) and 0.001 (*P+T_0.001*), frequentist LASSO regression model (*lassosum*)^18,19^, nonparametric Bayesian Dirichlet process regression model (*SDPR*)^20,21^, and Bayesian multiple linear regression model with continuous shrinkage prior (*PRS-CS*)^22^. These PRS models will estimate five sets of pQTL weights 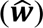 for each protein-coding gene per tissue type.

In Stage II, OTTERS first uses these five sets of pQTL weights 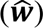 from Stage I as variant weights (*Equation 3*) to test gene-based association with respect to the phenotype in the summary-level GWAS test data. The test statistic can be written as

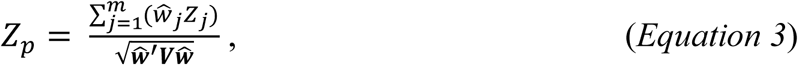

where *Z*_*j*_ denotes the single-variant Z-score test statistic in GWAS summary data for the *j*^*th*^ genetic variant, and ***V*** denotes the genotype correlation matrix that could be obtained from an external reference panel of the same ancestry as the test GWAS data^8^. Such gene-based association test has been shown to be equivalent as testing the association between predicted genetically regulated protein abundances and the phenotype in the GWAS test data^7,8,23^.

Since the performance of a PRS model depends on the unknown genetic architecture of protein abundances, OTTERS aggregates the PWAS p-values based on all five PRS models by using the aggregated Cauchy association test (ACAT)^24^. An omnibus test p-value is derived for each protein-coding gene, which is then used to identify significant PWAS risk genes^11^.

### Applying OTTERS to conduct PWAS of AD dementia

We first applied OTTERS^11^ to estimate pQTL weights from the recently released summary pQTL data of brain (n=380), CSF (n=835), and plasma (n=529)^12^. These summary pQTL data were generated by using proteomic data of individuals with AD and cognitively normal individuals of European ancestry profiled from an aptamer-based platform^25^. These summary pQTL data were generated for 1079 proteins in brain, 731 proteins in CSF, and 931 proteins in plasma, and ∼14M genetic variants with minor allele frequency (MAF) ≥ 2%. Linear regression models with protein abundances as the response variable, genotype of a single genetic variant as the test covariate, and additional adjusting covariates of age, sex, first two genotype principal components factors, and genotype platform, were used to generate the summary pQTL data. Since the summary pQTL data of these three tissues were generated by using samples of European ancestry, the LD information obtained from the whole genome sequencing data of European samples from the ROS/MAP study^9^ was used to estimate pQTL weights, along with standardized marginal pQTL effect sizes and sample sizes.

For each protein in these three tissue types, we obtained five sets of estimated pQTL weights by five PRS models as implemented by OTTERS, which were used to conduct PWAS analyses with the recent GWAS summary data (n=∼789K) of AD dementia^3^. The GWAS summary data of AD dementia^3^ were generated by meta-analysis, including clinically diagnosed AD cases and proxy AD and related dementia (proxy-ADD) cases from the UK Biobank (UKBB), resulting in a total of approximately 39,106 clinically diagnosed AD cases, 46,828 proxy-ADD cases, and 401,577 controls. By combining PWAS p-values based on five PRS models, the omnibus OTTERS p-values were obtained for each available protein in each of the three tissues. We corrected the omnibus OTTERS p-values per tissue by using the genomic control factor^26^ to ensure that the median observed OTTERS p-value was adjusted to the expected value of 0.5 under the null hypothesis. We then used the adjusted nominal OTTERS p-values to calculate the false discovery rates (FDR, i.e., q-values) per tissue. Genes with q-values < 0.05 were identified as significant PWAS risk genes for AD dementia in the corresponding tissue.

### Causal effects of PWAS risk genes by PMR-Egger

The two-stage PWAS framework does not distinguish genetic effects on phenotype that are mediated through genetically regulated protein abundances (i.e., causal effects or vertical pleiotropy effects), from shared genetic effects on protein abundances and phenotypes that are not mediated through protein abundances (i.e., horizontal pleiotropy effects). We further assessed the mediated causal effects of our identified significant PWAS risk genes by using the probabilistic Mendelian randomization (PMR-Egger) tool^27^. PMR-Egger can assess causal mediated genetic effects while controlling for horizontal pleiotropy effects by using summary pQTL and GWAS data. The reference LD derived from the ROS/MAP WGS data was also used for implementing PMR-Egger.

### PPI network and enrichment analyses by STRING

The STRING^28–30^ webtool integrates public data sources of protein interaction and analyzes the protein-protein interaction (PPI) network connectivity of proteins. Protein-protein edges represent the functional association, colored with six different connections –– curated databases, experiments, text mining, co-expression, gene co-occurrence and protein homology. Gene co-occurrence association predictions are based on whole-genome comparisons. The STRING^28^ webtool also provides gene enrichment analysis with respect to Gene Ontologies (GO)^31^ annotations. Enrichment analysis aims to detect GO terms and pathways that are significantly enriched with genes in the network versus random genes. The enrichment strength is provided along with FDR, which indicates the ratio between the number of proteins in the network that are annotated with a term and the number of proteins that expected to be annotated with this term in a random network of the same size. In this study, we utilized the STRING webtool to conduct PPI network and enrichment analyses with the list of PWAS risk genes identified by OTTERS in all three tissues.

### TWAS of AD dementia

We also conducted TWAS of AD dementia by using the same GWAS summary data of AD dementia^3^ and the reference transcriptomic data of 931 DLPFC samples from the ROS/MAP studies^9^. ROS and MAP are two ongoing longitudinal studies of aging and Alzheimer’s disease. Both studies enroll participants without known dementia, who agree to annual clinical evaluations and brain donation upon death. Transcriptomic data of 931 DLPFC samples from the ROS/MAP studies were profiled by RNA-sequencing. Gene expression data were quantified by transcripts per million (TPM), log2 transformed, and adjusted for confounding covariates, including age, sex, postmortem interval, batch effects, and 20 PEER factors. Quality-controlled ROS/MAP WGS data were used for TWAS, where common genetic variants with minor allele frequency (MAF) > 1% and Hardy-Weinberg p-value > 10^-5^ were analyzed. Cis-genetic variants within 1Mb of each gene’s flanking regions were used in the gene expression imputation models as predictors. Reference LD derived from ROS/MAP WGS samples of European ancestry were used in the TWAS.

For each gene, three TWAS tools including TIGAR, PrediXcan, and FUSION were utilized for estimating eQTL weights in Stage I. The Stage II TWAS results of AD dementia were obtained by using the eQTL weights estimated by each of the TWAS tool, which were combined by the ACAT method^10^. The combined omnibus TWAS p-values were adjusted for the genomic control factor^26^, and then used to derive FDR q-values. Significant TWAS risk genes were identified with q-values < 0.05.

## Results

### PWAS results of AD dementia by OTTERS

By applying OTTERS^11^ to the summary pQTL reference data of three tissues (brain, CSF and plasma)^5^ and recent large-scale GWAS summary data of AD dementia, we obtained PWAS p-values with pQTL weights estimated by five complimentary PRS models (see Methods). Moderate inflation was observed in the Quantile-Quantile (Q-Q) plots of these proteome-wide p-values in all three tissues (**Fig. S1-S3**). Omnibus OTTERS p-values were obtained by combining the PWAS p-values across all 5 PRS methods^24^, and then were adjusted by the genomic control factor^26^. The adjusted OTTERS p-values were used to calculate FDR q-values to account for multiple testing. We identified 30 PWAS significant risk genes of AD dementia with OTTERS FDR q-value < 0.05, including 11, 9, and 16 genes respectively detected in brain, CSF, and plasma tissues (**Fig. 1**; **Table 1**), with 4 detected in at least two tissues, and gene *MAPK3* detected in all three tissues.

**Figure 1.**
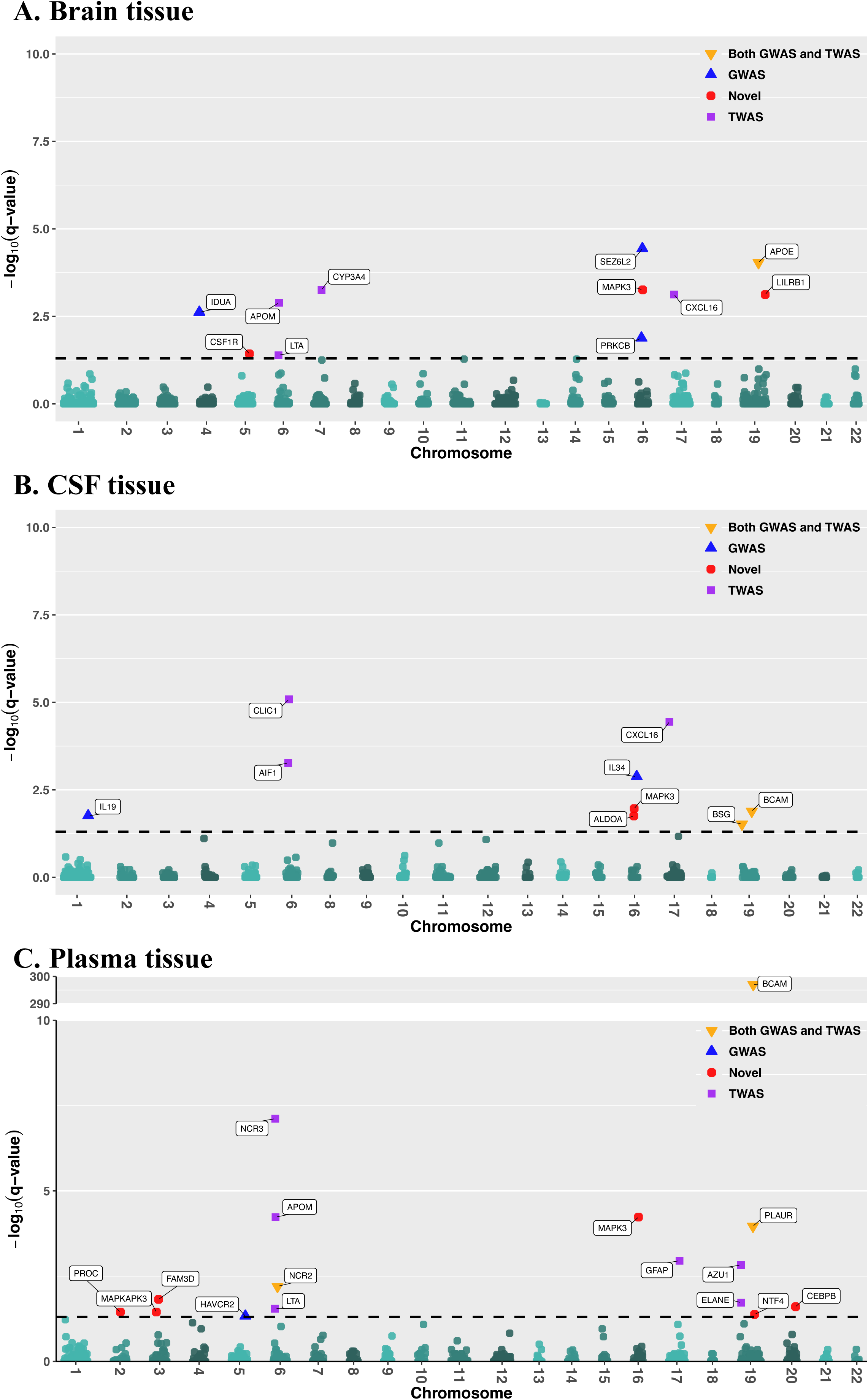
Manhattan plots of PWAS results (FDR q-values) of AD dementia by OTTERS in Brain (A), CSF (B), and Plasma (C) tissues. The −log10(q-values) were plotted on the y-axis, and −log10(0.05) was plotted as the dashed horizontal line. Independent significant genes are labeled. Yellow triangles: genes identified by previous GWAS and TWAS; Blue triangles: genes identified by previous GWAS; Purple square: genes identified by previous TWAS; Red circle: novel findings.

As shown in **Table 1**, we found 11 out of 30 PWAS significant genes (labeled in **Fig. 1**), such as *BCAM*^32^*, APOE*^33^*, IL19*^34^*, and IL34*^35^, were detected by previous GWAS of AD or AD pathological hallmarks as shown in GWAS Catalog^36^. We also found 2 of these PWAS risk genes (labeled in **Fig. 1**) identified in brain (*APOE, APOM*), 2 in CSF *(AIF1, BCAM*), and 2 in plasma (*APOM, BCAM*) were identified by previous PWAS of AD dementia using individual-level proteomic data profiled from DLPFC tissue of the ROS/MAP cohorts^10^.

Additionally, our analysis identified 9 novel PWAS risk genes (labeled in **Fig. 1**) that are not previously associated with AD dementia by GWAS, TWAS or PWAS –– *CSF1R, MAPK3* and *LILRB1* in brain; *ALDOA and MAPK3* in CSF; *PROC, MAPKAPK3, FAM3D, MAPK3, NTF4,* and *CEBPB* in plasma. AD relevant biological functions were reported for these novel findings. Especially, *CSF1R* that encodes the tyrosine kinase receptor *CSF1R* was associated with microglial homeostasis and neuronal survival, implicating its dysfunction in increasing risks for AD^37^. *LILRB1* that encodes a receptor for soluble β-amyloid (Aβ) oligomers was found as a potential contributor to synaptic loss and cognitive deficits in AD^38^. Alterations in *ALDOA*, implicated in oxidative neurotoxicity, were found to contribute to the pathogenesis of Alzheimer’s disease^39^. *PROC* is involved in the neurodegenerative process, showing an inverse association with incident dementia^40^. The stress-responsive kinase *MAPKAPK3*, a downstream target of *MAPK14*, was implicated in the regulation of autophagy and might play a role in AD pathology by modulating the degradation of proteins involved in amyloid plaque formation^41^. *FAM3D* was found involved in neuroinflammatory pathways and microglial responses in early AD stage^42^. The neurotrophic factor *NTF4*, was shown to play a role in early neurodegenerative changes and cognitive decline in AD^43^. The transcription factor *C/EBPβ,* encoded by *CEBPB*, has been implicated in the regulation of *APOE* expression^44^. The gene *MAPK3* detected in all three tissues is involved in immune system processes, which was found to be activated in AD brains and involved in pathogenesis of AD including tau phosphorylation and amyloid deposition^45^.

### Comparison of different PRS models

As described in the Methods section, OTTERS leverages all 5 complimentary PRS model (*P+T_0.001, P+T_0.05*, *lassosum*, *SDPR*, and *PRS-CS*) to account for complex genetic architecture underlying the protein abundance quantitative traits, thus improving the PWAS power for studying complex diseases. Here, we compared the omnibus OTTERS p-values to the PWAS p-values obtained by each individual PRS methods (**Table 1**). As pointed out by the OTTERS paper^11^, we found that all individual PRS models contributed to the omnibus OTTERS results. For example, the well-known AD risk gene *APOE* in brain was detected by all PRS models except *P+T_0.001,* and gene *BCAM* in CSF that is proximate to *APOE* was detected by *P+T_0.05, PRS-CS,* and *SDPR*. To investigate how individual PRS models contribute to the final OTTERS results, we plotted the pQTL weights estimated by all five PRS models for three example PWAS risk genes (*APOE*, *BSG*, and *PROC*) that were respectively detected in brain, CSF, and plasma tissues by OTTERS (**Fig. 2**). We plotted these pQTL weights with colors corresponding to −*log*10 (GWAS p-values).

**Figure 2.**
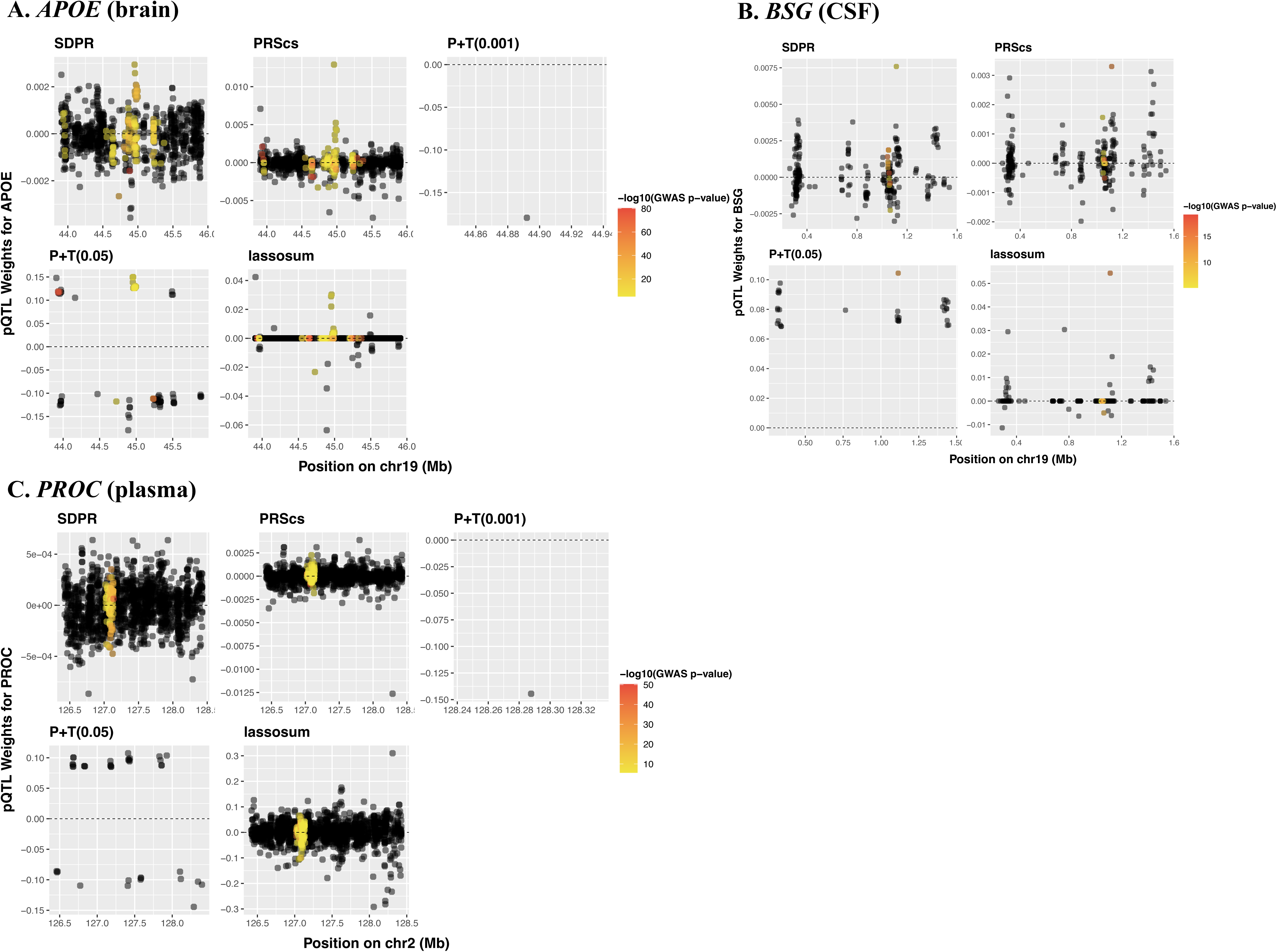
Scatter plots of pQTL weights of example PWAS risk genes of *APOE* in brain, *BSG* in CSF, and *PROC* in plasma that were estimated by individual PRS models. The pQTL weights were plotted in the y-axis for all genetic variants in the test gene region and color-coded with respect to −log10 (GWAS p-value). Test SNPs with GWAS p-value <10^−5^were colored.

In **Fig. 2A**, we showed pQTL weights for *APOE* in brain, which was found significant by *lassosum* (p-value = 1.23e-10), *P+T_0.05* (p-value = 1.23e-05), *PRS_CS* (p-value = 1.35e-05), and *SDPR* (p-value = 8.45e-07). We found that the significance of gene *APOE* was driven by GWAS significant SNPs located in *TOMM40* and *NECTIN2*, with non-zero pQTL weights estimated by these PRS methods. Most genetic variants with “significant” GWAS p-values were excluded by *P+T_0.01* due to their pQTL p-values > 0.001. Interestingly, genes *TOMM40* and *NECTIN2* are proximal to *APOE. TOMM40* is a known risk gene of AD dementia^1^ and has been associated with family history of AD^46^. *NECTIN2* was previously reported as a GWAS risk gene for beta-amyloid 1-42^35^, low-density lipoprotein cholesterol level interaction, and short total sleep time^47^.

In **Fig. 2B**, we showed pQTL weights of gene *BSG* in CSF, which was detected by *lassosum* (p-value =1.54e-05), *PRS-CS* (p-value = 3.07e-04), and *SDPR* (p-value = 9.96e-05). Compared to the *P+T* models, these PRS methods all estimated non-zero pQTL weights for more GWAS significant SNPs in the test region. In **Fig 2C**, we showed pQTL weights of gene *PROC* in plasma, which was only detected by *PRS-CS* (p-value = 4.31e-05) and *SDPR* (p-value = 9.10e-04). *lassosum* estimated pQTL weights with higher magnitude for “non-significant” GWAS SNPs (GWAS p-values > 1e-5, black dots) in the test region, and most “significant” GWAS SNPs were filtered out by the *P+T* models due to their pQTL p-values >0.001.

Additionally, we plotted the pQTL weights of another 3 significant PWAS risk genes –– *APOM* in brain (**Fig. S4)***, AIF1* in CSF **(Fig. S5**), and *NCR2* in plasma **(Fig. S6)**. *APOM* (p-value=2.42e-08 in brain) and *AIF1* (p-value = 1.22e-08 in CSF) were found significant only by *lassosum*, which estimated pQTL weights in relatively higher magnitude for “significant” GWAS SNPs. *NCR2* (in plasma) were found significant by all PRS methods except *P+T_0.001* that estimated non-zero pQTL weights for “significant” GWAS SNPs.

Overall, these plots of pQTL weights demonstrated that significant PWAS risk genes were mainly driven by test genetic variants that had non-zero pQTL weights with relatively large magnitudes and had “significant” GWAS p-values (e.g., <10e-05). These pQTL weight plots also showed that OTTERS leveraged the strength of all complementary PRS methods to achieve higher power.

### Mediated causal effects of PWAS risk genes

We employed the probabilistic Mendelian Randomization tool, PMR-Egger^27^, to assess if the genetic effects of these 30 PWAS risk genes were mediated through genetically regulated protein abundances and causal for AD dementia, while accounting for possible horizontal pleiotropy effects (Methods). As shown in **Table 1**, we found that 4 out of 11 (36.4%) PWAS risk genes in brain, 6 out of 9 (66.7%) PWAS risk genes in CSF, 4 out of 16 (25.0%) PWAS risk genes in plasma tissues had significant mediated causal genetic effects with FDR < 0.05 without significant horizontal pleiotropy effects. Additionally, we found 3 PWAS risk genes in plasma with significant mediated causal genetic effects that also had significant horizontal pleiotropy effects with FDR < 0.05. In summary, by PMR-Egger, we validated causal genetic effects that were mediated through genetically regulated protein abundances for 13 (43.3%) out of these 30 PWAS risk genes in brain, CSF, and plasma tissues by OTTERS.

### PPI network and enrichment analyses

By using the STRING^28^ webtool (Methods), we found 25 out of these 30 significant PWAS risk genes identified by OTTERS were interconnected in a network involving the well-known AD risk gene *APOE* (**Fig. 3**). The edges of the network were colored according to the protein-protein interactions (PPI) based on different data sources. The STRING webtool also provided gene enrichment analyses results with these PWAS risk genes (**Fig. 3**).

**Figure 3.**
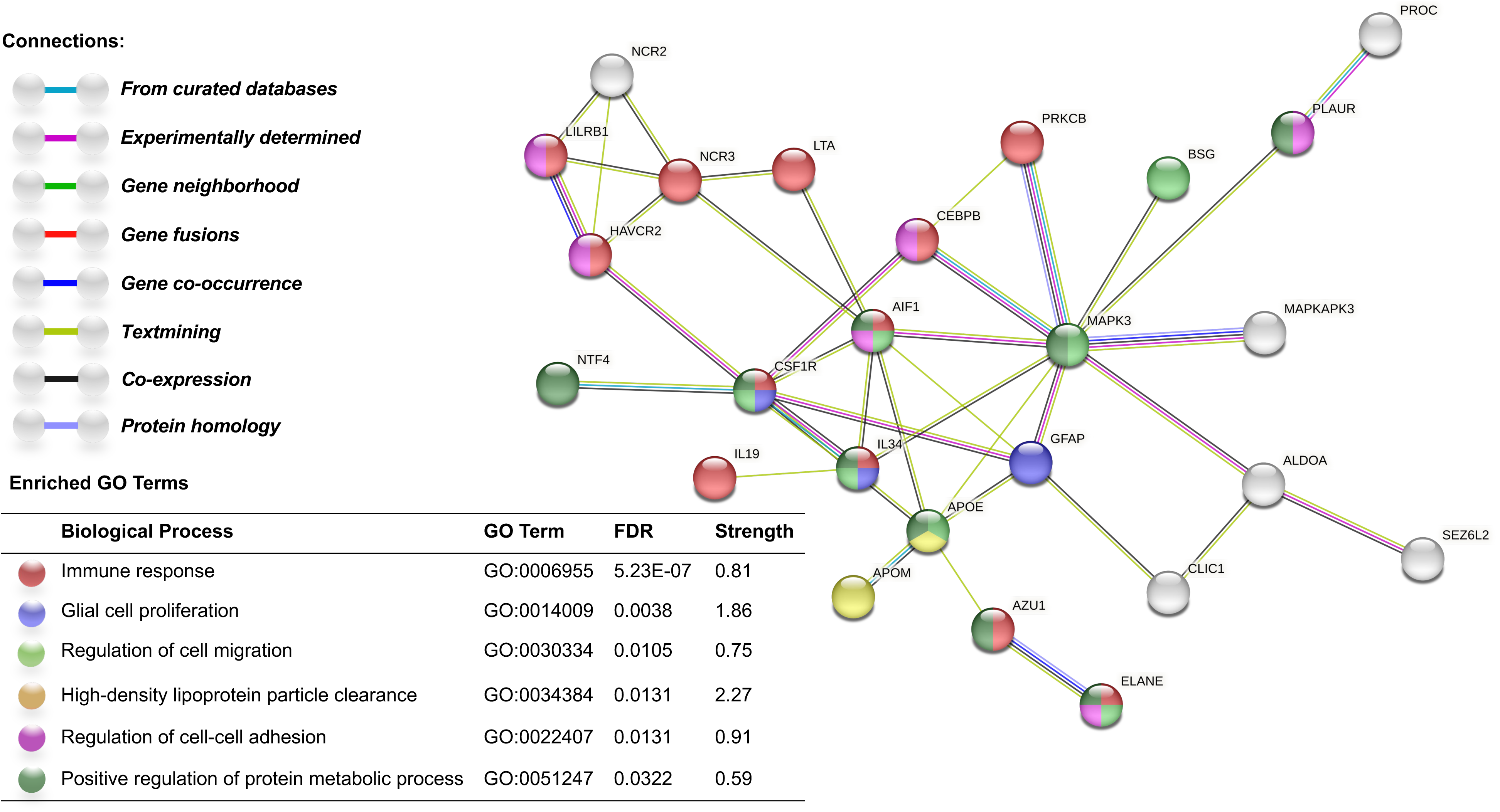
PPI network and enrichment analyses results with 25 PWAS risk genes of AD dementia by STRING. Edges represent physical PPI, with different colors representing different sources of connection evidence. Node colors represent different enriched GO terms with FDR < 0.05, 6 GO terms with most significant FDR q-value were colored.

We found that our identified 30 PWAS risk genes were enriched in gene ontology (GO) pathways of immune response (*AZU1, CSF1R, CXCL16, CEBPB, HAVCR2, LILRB1, NCR3, AIF1, IL34, LTA, ELANE, PRKCB, IL19*) with enrichment FDR = 5.23e-07 and strength = 0.81, glial cell proliferation (*CSF1R, CEBPB, IL34*) with enrichment FDR = 0.0038 and strength = 1.86, regulation of cell migration (*APOE, MAPK3, CSF1R, CXCL16, BSG, AIF1, IL34, ELANE*) with enrichment FDR = 0.0105 and strength = 0.75, regulation of cell-cell adhesion (*CEBPB, HAVCR2, LILRB1, PLAUR, AIF1, ELANE*) with enrichment FDR = 0.0131 and strength = 2.27, high-density lipoprotein particle clearance (*APOE, APOM*) with enrichment FDR = 0.0131 and strength = 0.91, as well as positive regulation of protein metabolic process (*AZU1, APOE, MAPK3, CSF1R, PLAUR, AIF1, IL34, ELANE, NTF4*) with enrichment FDR = 0.0322 and strength = 0.59. Also, we found that 14 of these PWAS risk genes were enriched in the Human Phenotype of blood protein measurement (e.g., *MAPK3, APOE, PRKCB*) with enrichment FDR = 0.0014 and strength = 0.71; and 15 of these were enriched in immune system (e.g., *IL34, NCR3, AZU1*) in the Reactome Pathway^48^, with enrichment FDR = 9.03e-05 and strength = 0.7.

Particularly, the detected PPI network showed that the novel PWAS risk genes identified by OTTERS were closely interconnected with known risk genes of AD dementia. For example, novel PWAS risk gene *CEBPB* was connected with the known GWAS and TWAS risk gene *PRKCB*^49^ and was in the GO pathway of immune response. Novel risk genes *CSF1R a*nd *NTF4* were found connected with the known GWAS risk gene of AD dementia *IL34*^50^ and were in the GO pathway of positive regulation of protein metabolic process.

### Compare to TWAS results of AD dementia

Next, we compared our PWAS findings to the TWAS findings by using individual-level reference transcriptomic data of DLPFC from the ROS/MAP studies and the same GWAS summary data of AD dementia. As described in Methods, we utilized the omnibus TWAS approach^10^ by using the ACAT method^24^ to combine the results obtained by individual TWAS tools of TIGAR/DPR, PrediXcan/Elastic-Net, and FUSION/BestModel.

The omnibus TWAS (TWAS-O) approach identified 113 significant risk genes of AD dementia with FDR q-value < 0.05 (**Fig. 4**), including a total of 17 independent significant genes (labeled in **Fig. 4**) that do not have shared genetic variants in their test regions (±1MB around the test transcription starting/termination sites of the protein-coding gene). These independent significant genes were curated as having no overlapped test genetic variants. Importantly, 18 out of our identified 30 PWAS risk genes were either identified by TWAS-O or located within the shared (1Mb) test regions of the risk genes identified by TWAS-O, including 7 out of 11 in brain, 6 out of 9 in CSF, and 9 out of 16 in plasma tissues.

**Figure 4.**
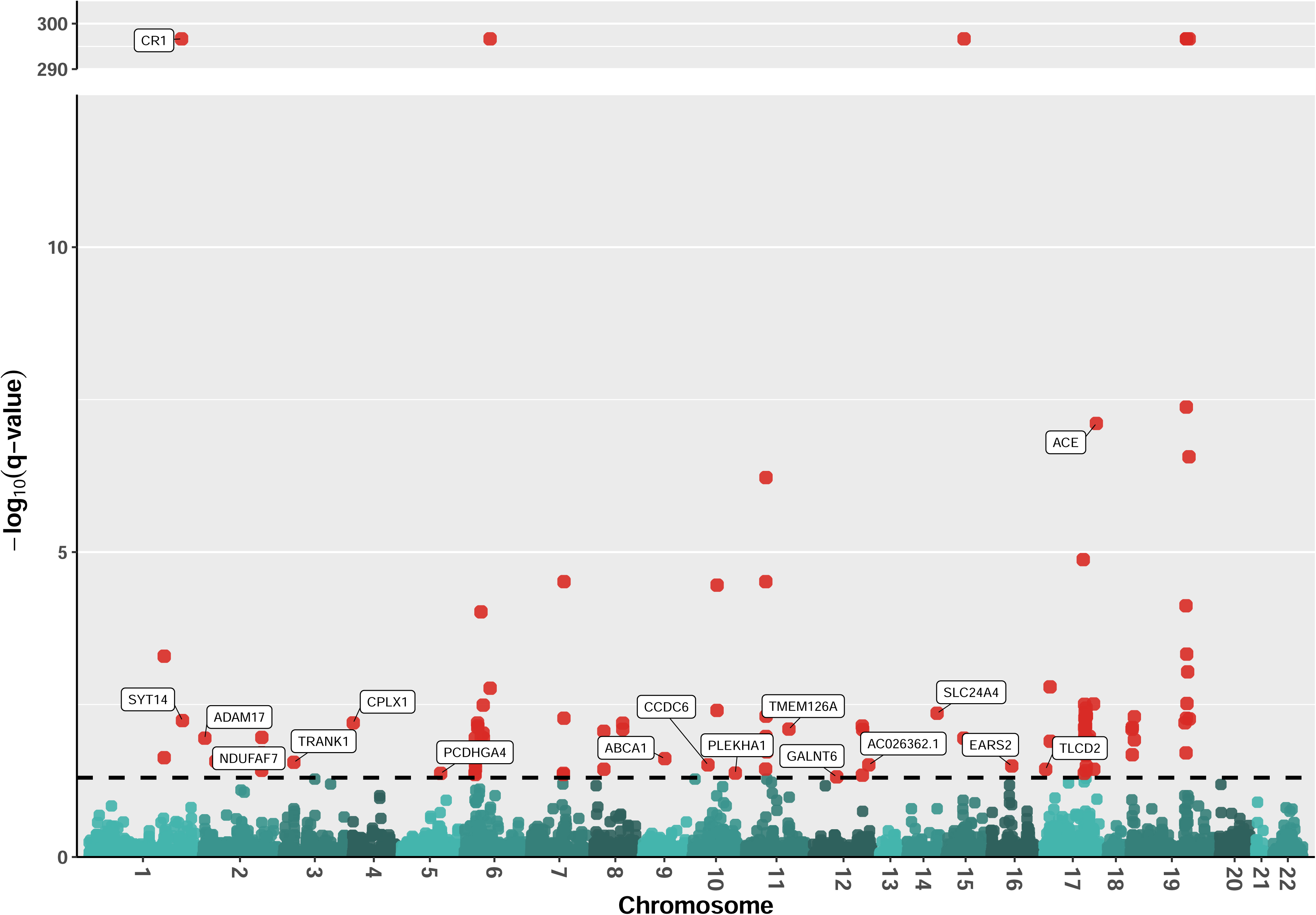
Manhattan plot of TWAS results of AD dementia in DLPFC tissue. The −log10(q-value) were plotted on the y-axis, and −log10(0.05) was plotted as the dashed horizontal line. Plotted TWAS q-values were obtained by the ACAT method, combining TWAS p-values obtained by TIGAR/DPR, PrediXcan/Elastic_Net, and FUSION/BestModel. Independent significant genes are labeled.

For example, the PWAS risk gene *IDUA* in brain has shared test genetic variants as the TWAS-O risk gene *CPLX1* (q-value = 6.31e-03), whose downregulation is likely to influence AD-associated neurodegeneration^51^. The PWAS risk gene *APOE* in brain has shared test genetic variants with 14 nearby TWAS-O risk genes on chromosome 19. PWAS risk genes *AIF1* and *CLIC1* in CSF have shared test genetic variants with the same set of 8 TWAS-O risk genes, which include *HLA-DRB1*, a known risk gene of late-onset Alzheimer’s disease (LOAD), particularly in APOE ε4 non-carrier population^52^. PWAS risk gene *BCAM* in both CSF and plasma tissues has overlapped test genetic variants with 18 TWAS-O risk genes, including *ZNF112, ZNF 221 and ZNF 233* that are Zinc finger protein genes known with pathophysiological role in neuro-related diseases and disorders^53^. PWAS risk gene *GFAP* in plasma has shared test genetic variants with 19 TWAS-O risk genes, including *MAPT* that encodes the tau protein with a critical role in neurodegeneration^54^.

## Discussion

We conducted PWAS of AD dementia with the recently released public data resources of summary pQTL data of brain, CSF, and plasma tissues^12^ as well as GWAS summary data of AD dementia, by employing our recently developed OTTERS^11^ tool. The distinction of our PWAS analyses from previous ones lies in the use of summary pQTL reference data from multiple tissues (brain, CSF, and plasma) related to neurodegenerative disorders. CSF and plasma are two bodily fluids believed to contain the richest source of biomarkers of AD and play important roles in research of AD pathology^55^. CSF surrounds the central nervous system (CNS) and is a highly representative and obtainable fluid for detecting brain pathologies. Blood plasma contains proteins that affect brain functions from the periphery, as well as proteins exported from the brain^55^. Especially, recent studies have shown that amyloid beta and phosphorylated tau presenting in CSF and plasma could be used as biomarkers for detecting AD dementia in early stages^13–16^. Therefore, our PWAS results leveraging proteomic data of plasma and CSF tissues in addition to brain tissue are expected to reveal additional important risk genes of AD, whose genetic effects are mediated through protein abundances in biofluids, thus providing valuable insights into future biomarker discovery of AD dementia.

We identified 30 PWAS risk genes whose genetic effects were potentially mediated through genetically regulated protein abundances, including 11 in brain, 9 in CSF and 16 in plasma tissues. We found OTTERS gained power by leveraging multiple complementary PRS models to estimate pQTL weights and by considering reference pQTL data of multiple tissues. Specifically, we showed that each PRS model made distinct and considerable contributions to the final omnibus PWAS results by OTTERS.

Previous studies have highlighted notable biological roles for our identified PWAS risk genes in three tissues, including significant enrichment in the GO biological pathways of immune system processes and lipoprotein metabolism. For example, PWAS risk gene *APOE* in brain is a known major risk factor for AD dementia^56^, which has a crucial function in the central nervous system^57^. The PWAS risk gene *APOM* in brain and plasma codes for an apolipoprotein associated with AD dementia and has been implicated in the lipid processing pathway^58^. The PWAS risk gene *AIF1* in CSF has been associated with the activation of microglia (a type of immune cell localized throughout the central nervous system)^59^, which is a key player in the response to central nervous disorders such as AD^60^.

Besides known AD risk genes, our PWAS also identified 9 novel risk genes that are not detected by previous GWAS, TWAS, or PWAS. Especially, 6 out of these 9 novel PWAS risk genes were connected with known AD risk genes in the PPI network. Novel PWAS risk genes *CSF1R, LILRB1,* and *CEBPB* are enriched in the pathway of immune response, along with previously known AD risk genes.

These previous findings, results from Mendelian Randomization analysis using by PMR-Egger, and PPI network analysis results collectively demonstrate the significance of our identified PWAS risk genes in three tissues. Further experimental studies about the functions of our findings are essential but out of the scope of this work.

PWAS analysis by the OTTERS tool still has its limitations. First, we only consider *cis*-pQTL within the ±1*Mb* region around the transcription start/termination sites of the corresponding protein-coding gene. Second, the two-stage PWAS cannot account for possible horizontal pleiotropy genetic effects (those directly affecting the phenotype of interest) when testing if the genetic effects are mediated through genetically regulated protein abundances. Although the PMR-Egger tool can account for horizontal pleiotropy genetic effects, the computation burden of the PMR-Egger tool impedes its application to testing the protein-coding genes of proteome-wide proteins (average 60 vs. 2 CPU minutes per protein-coding gene by OTTERS). Thus, we only applied the PMR-Egger^27^ tool to our identified 30 PWAS risk genes.

## Conclusions

In conclusion, we presented the first PWAS analysis of AD dementia utilizing the summary pQTL reference data of multiple tissues related to neurodegenerative disorders. Our identified PWAS risk genes provide candidate proteins in biofluids such as CSF and plasma and brain tissues for follow-up functional experiments and targeted therapeutic development of AD dementia. Previous studies, validation results from Mendelian Randomization analysis by PMR-Egger, and PPI network analysis all demonstrated the significance of our identified PWAS risk genes in these three tissues. Further experimental studies about the functions of our findings are essential. Additionally, this study showed the practical usefulness of the OTTERS tool in leveraging publicly available summary pQTL data and GWAS data resources to conduct PWAS of complex diseases.

## Data Availability

All data produced in the present study are available upon reasonable request to the authors

## List of Abbreviations

PWAS: Proteome-wide association study
GWAS: Genome-wide association study
AD: Alzheimer’s disease
pQTL: Protein quantitative trait loci
CSF: Cerebrospinal fluid
PRS: Polygenic risk score
ACAT: Aggregated Cauchy association test
PPI: Protein-protein interaction
PMR-Egger: Probabilistic Mendelian randomization Egger
TWAS: Transcriptome-wide association study
DLPFC: Dorsolateral prefrontal cortex
WGS: Whole genome sequencing
proxy-ADD: proxy AD and related dementia
UKBB: UK Biobank
LD: Linkage disequilibrium
FDR: False discovery rate
ROS/MAP: Religious Orders Study and Memory and Aging Project
GO: Gene Ontologies
MAF: Minor allele frequency
TPM: Transcripts per million
PEER: Probabilistic estimation of expression residuals

## Declarations

### Ethics approval and consent to participate

Summary-level pQTL data of brain, CSF, and plasma, and summary GWAS data of AD dementia were used for conducting PWAS in this study. These data are de-identified and publicly available, requiring no ethics approval. All de-identified omics data are not considered as human data per NIH guidelines.

All ROSMAP participants enrolled without known dementia and agreed to detailed clinical evaluation and brain donation at death [https://pubmed.ncbi.nlm.nih.gov/29865057/]. Both studies were approved by an Institutional Review Board of Rush University Medical Center (ROS IRB# L91020181, MAP IRB# L86121802). Both studies were conducted according to the principles expressed in the Declaration of Helsinki. Each participant signed an informed consent, Anatomic Gift Act, and an RADC Repository consent (IRB# L99032481) allowing her data and biospecimens to be repurposed. ROS/MAP transcriptomic and whole genome sequencing data used for TWAS in this study were shared with a data use agreement.

### Consent for publication

Not applicable.

### Availability of data and materials

GWAS summary data of AD dementia is available from^2^. Summary-level pQTL data of brain, CSF, and plasma tissues can be accessed by emailing to set up an FTP transfer of the data. Omics data from ROS/MAP cohorts are available with approved access from https://doi.org/10.7303/syn10901595. OTTERS tool is available from https://github.com/daiqile96/OTTERS. PMR tool is available from https://github.com/yuanzhongshang/PMR. The code used in this study for conducting PWAS of AD dementia are available from GitHub https://github.com/tingyhu45/PWAS_OTTERS. Trained pQTL weights by five PRS methods and our PWAS summary data will be deposited to SYNAPSE once this work is accepted. ROSMAP data can be requested at www.radc.rush.edu and www.synapse.org.

### Authors’ contributions

T.H. and Q.L. conducted data analysis and drafted manuscript. Q.D. and R.L.P. participated in data analysis and data interpretation, and revised manuscript. M.P.E. supervised data analysis, data interpretation, and revised manuscript. J.Y. conceived this study, supervised data analysis, data interpretation, and revised manuscript.

