## Supplemental Figure 1-7 for "Proteome-wide association studies using summary pQTL data of three tissues identified 30 risk genes of Alzheimer’s disease dementia"

**Table 1: Significant PWAS risk genes identified by OTTERS in brain tissue.** All PWAS P-values are adjusted by genomic control factor. P-values by individual PRS models with significant FDR<0.05 are bold. NA p-values by P+T models that are shown as “-” in the table are because all test SNPs have pQTL p-values > the corresponding threshold of 0.001 or 0.05.

| **Gene** | **CHR** | **PWAS P-values** | | | | | |
| --- | --- | --- | --- | --- | --- | --- | --- |
|  |  | **OTTERS** | **lassosum** | **P+T_0.001** | **P+T_0.05** | **PRS-CS** | **SDPR** |
| IDUA*^a,b^* | 4 | 3.57e-07 | **7.88e-08** | **1.40e-03** | **1.04e-02** | **9.59e-07** | **3.69e-06** |
| CSF1R | 5 | 2.27e-05 | **6.17e-06** | **5.94e-04** | **6.74e-04** | **2.40e-05** | **7.23e-05** |
| LTA*^b^* | 6 | 2.94e-05 | 8.43e-01 | - | **8.10e-06** | **1.53e-03** | **8.34e-05** |
| APOM*^b,c^* | 6 | 1.21e-07 | **2.42e-08** | 9.37e-01 | 3.01e-01 | 5.67e-01 | 2.36e-01 |
| CYP3A4*^b,d^* | 7 | 1.27e-08 | 6.77e-01 | **2.54e-09** | **5.30e-04** | 2.52e-02 | 1.14e-01 |
| PRKCB*^a,b^* | 16 | 4.59e-06 | 5.32e-01 | 9.97e-01 | **9.18e-07** | 1.77e-02 | 3.28e-02 |
| SEZ6L2*^a^* | 16 | 5.09e-11 | 6.77e-02 | - | 5.18e-02 | **3.25e-10** | **1.33e-11** |
| MAPK3*^d^* | 16 | 1.66e-08 | 1.41e-02 | - | **4.15e-09** | **1.10e-02** | **8.89e-04** |
| CXCL16*^b^* | 17 | 4.54e-08 | 3.69e-01 | 5.84e-01 | **9.21e-09** | **1.98e-05** | **6.57e-07** |
| APOE*^a,b,c,d^* | 19 | 4.91e-10 | **1.23e-10** | - | **1.23e-05** | **1.35e-05** | **8.45e-07** |
| LILRB1*^d^* | 19 | 4.25e-08 | **2.15e-08** | **9.85e-07** | **6.70e-07** | **1.69e-08** | **1.03e-07** |

*a* Known GWAS risk genes of AD.

*b* Known TWAS risk genes of AD, or with test regions overlapped with known TWAS risk genes.

*c* Known PWAS risk genes of AD, or with test regions overlapped with known PWAS risk genes.

*d* Has significant causal genetic effects by PMR-Egger.

| **Gene** | **CHR** | **PWAS P-values** | | | | | |
| --- | --- | --- | --- | --- | --- | --- | --- |
|  |  | **OTTERS** | **lassosum** | **P+T_0.001** | **P+T_0.05** | **PRS-CS** | **SDPR** |
| IL19*^a,b,d^* | 1 | 1.94e-05 | 1.21e-01 | - | 7.28e-02 | **5.10e-06** | **1.00e-04** |
| AIF1*^b,c,d^* | 6 | 6.12e-08 | **1.22e-08** | 1.71e-01 | 1.85e-02 | 4.97e-02 | 8.02e-01 |
| CLIC1*^b^* | 6 | 5.53e-11 | **2.21e-11** | - | **2.05e-04** | **4.03e-11** | **4.69e-10** |
| ALDOA*^d^* | 16 | 2.19e-05 | **5.67e-06** | - | **2.09e-04** | **4.75e-03** | **8.95e-04** |
| MAPK3 | 16 | 6.24e-06 | **4.10e-06** | - | - | **1.30e-05** | - |
| IL34*^a,d^* | 16 | 2.93e-07 | **9.65e-07** | **1.01e-07** | **3.29e-07** | **5.84e-07** | **7.02e-07** |
| CXCL16*^b^* | 17 | 9.90e-10 | **7.55e-03** | - | **2.44e-07** | **2.94e-10** | **1.58e-09** |
| BSG*^a,b,d^* | 19 | 5.12e-05 | **1.54e-05** | - | 2.31e-02 | **3.07e-04** | **9.96e-05** |
| BCAM*^a,b,c,d^* | 19 | 9.88e-06 | 2.20e-01 | - | **2.49e-06** | **2.96e-04** | 2.35e-01 |

**Table 2: Significant PWAS risk gene identified by OTTERS in CSF tissue.** All PWAS P-values are adjusted by genomic control factor. P-values by individual PRS models with significant FDR<0.05 are bold. NA p-values by P+T methods that are shown as “-” in the table are because all test SNPs have pQTL p-values > the corresponding threshold of 0.001 or 0.05. For MAPK3, pQTL weights were not trained for SDPR model.

*a* Known GWAS risk genes of AD.

*b* Known TWAS risk genes of AD, or with test regions overlapped with known TWAS risk genes.

*c* Known PWAS risk genes of AD, or with test regions overlapped with known PWAS risk genes.

*d* Has significant causal genetic effects by PMR-Egger.

**Table 3: Significant PWAS risk gene identified by OTTERS in plasma tissue.** All PWAS P-values are adjusted by genomic control factor. P-values by individual PRS models with significant FDR<0.05 are bold. NA p-values by P+T models that are shown as “-” in the table are because all test SNPs have pQTL p-values > the corresponding threshold of 0.001 or 0.05.

| **Gene** | **CHR** | **PWAS P-values** | | | | | | |
| --- | --- | --- | --- | --- | --- | --- | --- | --- |
|  |  | **OTTERS** | **lassosum** | | **P+T_0.001** | **P+T_0.05** | **PRS-CS** | **SDPR** |
| PROC | 2 | 2.06e-04 | | 2.06e-02 | 8.77e-01 | 1.78e-01 | **4.31e-05** | **9.10e-04** |
| MAPKAPK3 | 3 | 2.07e-04 | | 1.73e-02 | - | 2.36e-01 | **5.93e-05** | **4.07e-04** |
| FAM3D | 3 | 4.58e-05 | | **3.38e-04** | - | - | **7.06e-04** | **1.64e-05** |
| HAVCR2*^a,d^* | 5 | 3.34e-04 | | **7.41e-05** | 5.85e-03 | **6.01e-03** | **2.26e-03** | **1.44e-03** |
| LTA*^b^* | 6 | 1.33e-04 | | **3.35e-05** | - | **4.51e-03** | 3.96e-02 | 6.79e-01 |
| NCR3*^b^* | 6 | 5.25e-12 | | **3.20e-12** | - | - | **4.49e-12** | **2.73e-11** |
| APOM*^b,c,d^* | 6 | 2.51e-08 | | **1.39e-05** | - | **8.79e-06** | **2.98e-08** | **7.96e-09** |
| NCR2*^a,b,d^* | 6 | 1.43e-05 | | **3.22e-05** | 1.63e-02 | **6.93e-06** | **8.85e-06** | **1.66e-05** |
| MAPK3 | 16 | 2.76e-08 | | **8.03e-05** | - | **4.44e-06** | **8.19e-07** | **6.96e-09** |
| GFAP*^b^* | 17 | 1.37e-06 | | **3.50e-07** | - | 1.70e-01 | **1.86e-05** | **1.35e-02** |
| AZU1*^b^* | 19 | 2.30e-06 | | 2.81e-02 | - | - | **7.67e-07** | 9.04e-01 |
| ELANE*^b^* | 19 | 6.72e-05 | | **2.36e-05** | - | - | 2.61e-02 | **4.65e-04** |
| PLAUR*^a,b,d^* | 19 | 7.29e-08 | | 2.10e-02 | - | **3.26e-03** | **1.82e-08** | **2.10e-05** |
| BCAM*^a,b,c^* | 19 | 1.78e-22 | | **3.24e-22** | - | **5.19e-23** | **1.59e-20** | **5.24e-17** |
| NTF4 | 19 | 2.67e-04 | | **8.97e-05** | - | - | 3.74e-02 | 2.29e-02 |
| CEBPB | 20 | 1.03e-04 | | **2.66e-05** | - | 8.00e-02 | **5.22e-03** | **9.84e-04** |

*a* Known GWAS risk genes of AD.

*b* Known TWAS risk genes of AD, or with test regions overlapped with known TWAS risk genes.

*c* Known PWAS risk genes of AD, or with test regions overlapped with known PWAS risk genes.

*d* Has significant causal genetic effects by PMR-Egger.
