## Supplementary material for "Proteome-wide association studies using summary pQTL data of three tissues identified 30 risk genes of Alzheimer’s disease dementia": Table 1-3

**Fig. S1. Quantile-quantile plots of PWAS results obtained by OTTERS with summary pQTL reference data of brain tissue.**

Genomic control factors were obtained as $\lambda=1.43$ for OTTERS omnibus PWAS p-values, $\lambda=1.45$ for PWAS p-values using pQTL weights estimated by lassosum, $\lambda=1.60$ by P+T_0.001, $\lambda=1.57$ by P+T_0.05, $\lambda=1.39$ by PRS-CS, $\lambda=1.35$ by SDPR. OTTERS omnibus PWAS p-values shown in A were adjusted by genomic control factor.

| **A. OTTERS** | **B. lassosum** | **C. P+T_0.001** |
| --- | --- | --- |
| 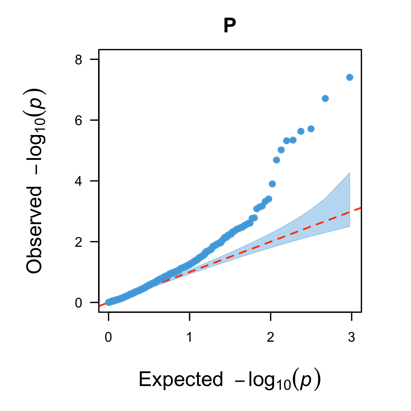 | 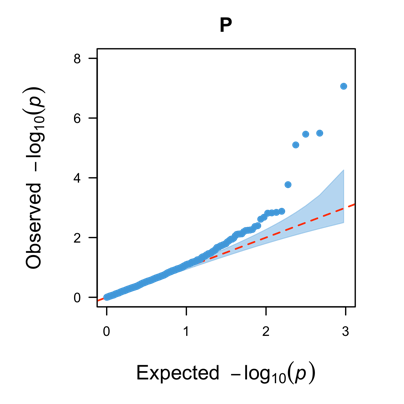 | 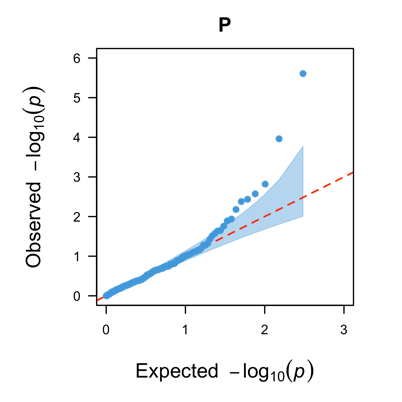 |
| **D. P+T_0.05** | **E. PRS-CS** | **F. SDPR** |
| 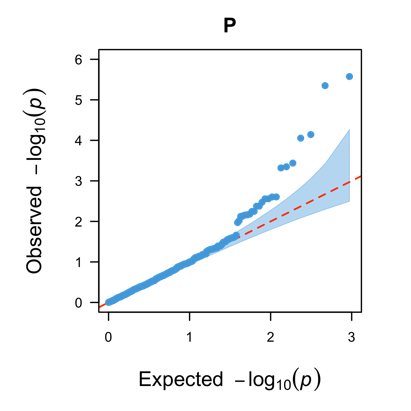 | 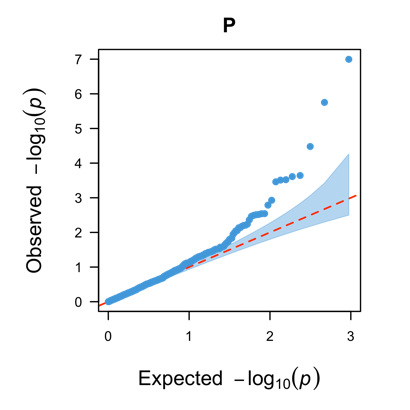 | 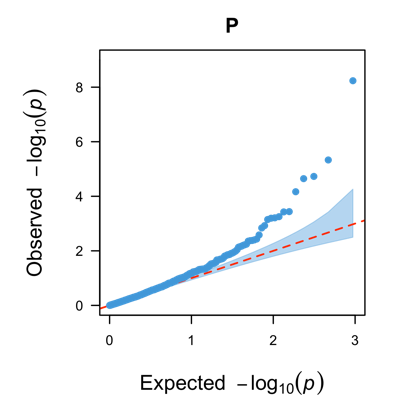 |

**Fig. S2. Quantile-quantile plots of PWAS results by OTTERS with summary pQTL reference data of CSF tissue.**

Genomic control factors were obtained as $\lambda=1.33$ for OTTERS omnibus PWAS p-values, $\lambda=1.40$ for PWAS p-values using pQTL weights estimated by lassosum, $\lambda=1.23$ by P+T_0.001, $\lambda=1.49$ by P+T_0.05, $\lambda=1.41$ by PRS-CS, $\lambda=1.30$ by SDPR. OTTERS omnibus PWAS p-values shown in A were adjusted by genomic control factor.

| **A. OTTERS** | **B. lassosum** | **C. P+T_0.001** |
| --- | --- | --- |
| 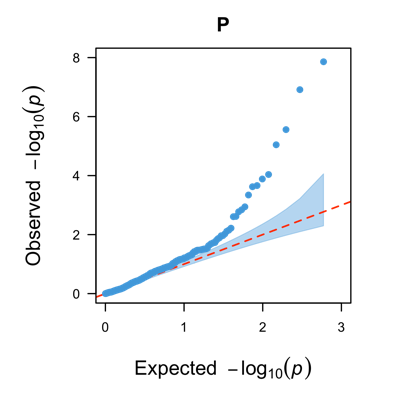 | 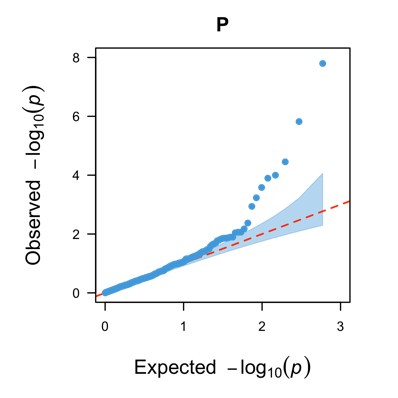 | 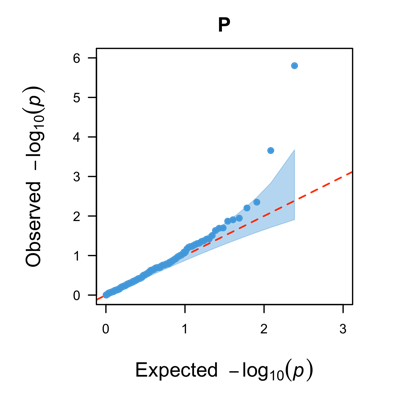 |
| **D. P+T_0.05** | **E. PRS-CS** | **F. SDPR** |
| 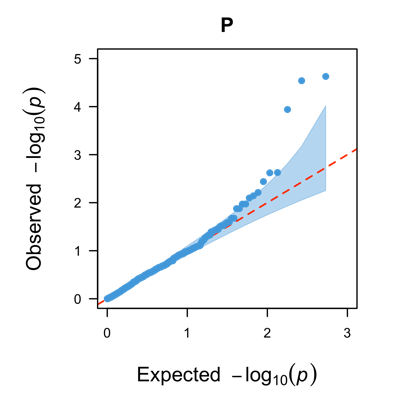 | 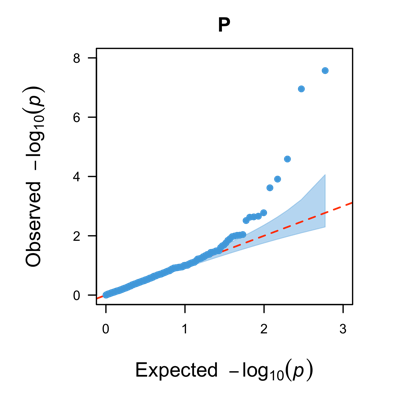 | 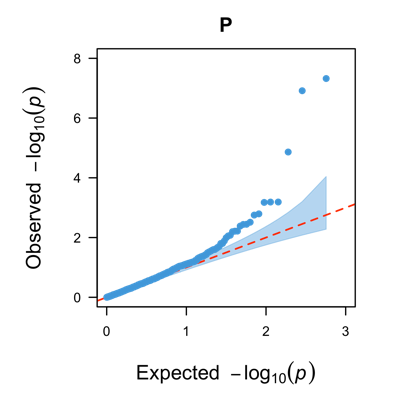 |

**Fig. S3. Quantile-quantile plots of PWAS results by OTTERS with summary pQTL reference data of plasma tissue.**

Genomic control factors were obtained as $\lambda=1.17$ for OTTERS omnibus PWAS p-values, $\lambda=1.51$ for PWAS p-values using pQTL weights estimated by lassosum, $\lambda=1.28$ by P+T (0.001), $\lambda=1.21$ by P+T (0.05), $\lambda=1.28$ by PRS-CS, $\lambda=1.38$ by SDPR. OTTERS omnibus PWAS p-values shown in A were adjusted by genomic control factor.

| **A. OTTERS** | **B. lassosum** | **C. P+T_0.001** |
| --- | --- | --- |
| 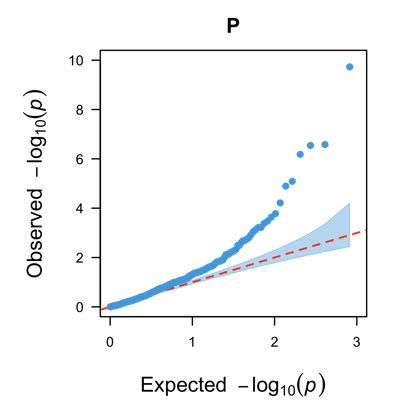 | 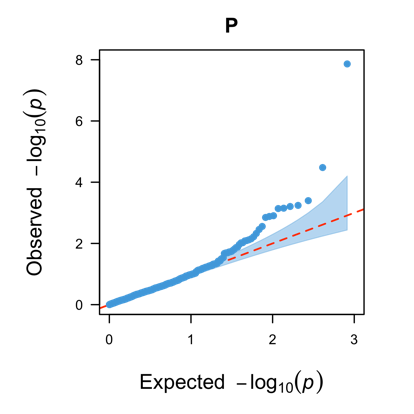 | 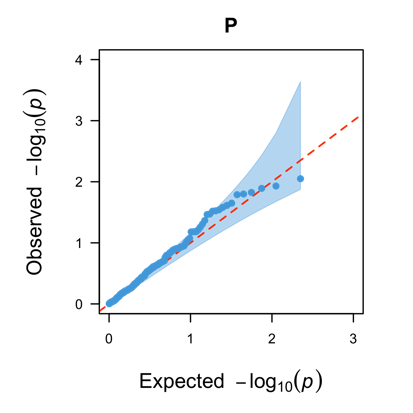 |
| **D. P+T_0.05** | **E. PRS-CS** | **F. SDPR** |
| 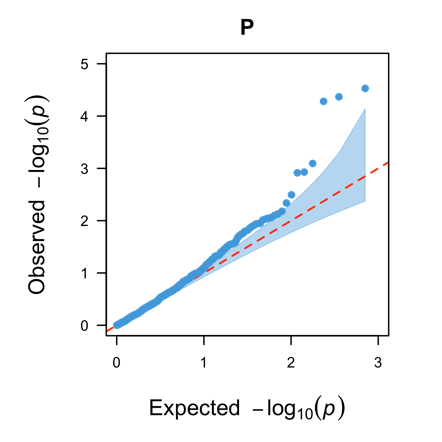 | 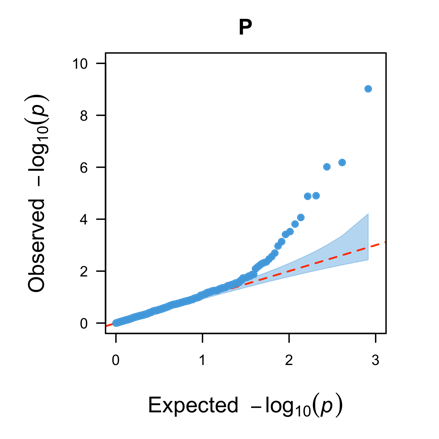 | 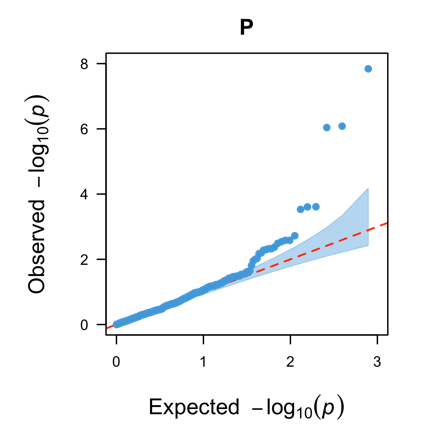 |

**Fig. S4: Scatter plots of pQTL weights estimated by individual PRS methods for PWAS risk gene *APOM* in brain tissue.** The pQTL weights were plotted in the y-axis for all test genetic variants in the test gene region, with color-coded with respect to -log10 (GWAS p-value). Test SNPs with GWAS p-value <${10}^{-5}$ were colored.

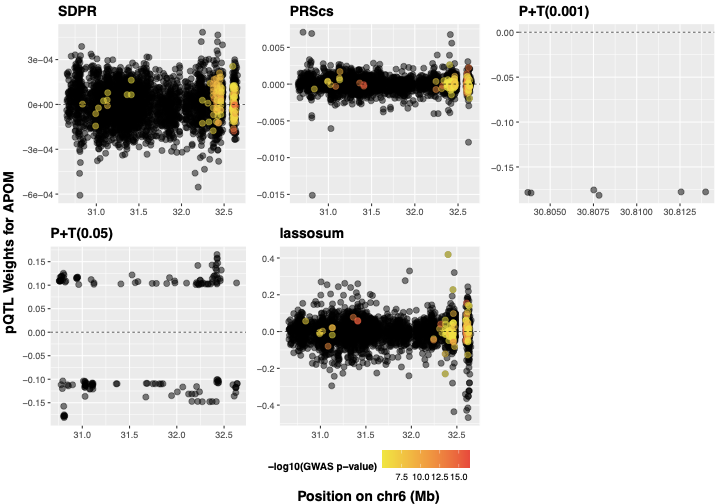

**Fig. S5: Scatter plots of pQTL weights estimated by individual PRS methods for PWAS risk gene *AIF1* in CSF tissue.** The pQTL weights were plotted in the y-axis for all test genetic variants in the test gene region, with color-coded with respect to -log10 (GWAS p-value). Test SNPs with GWAS p-value <${10}^{-5}$ were colored.

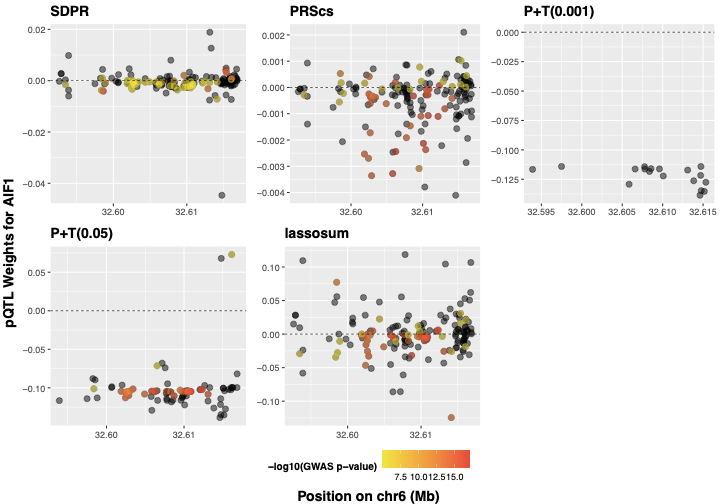

**Fig. S6: Scatter plots of pQTL weights estimated by individual PRS methods for PWAS risk *NCR2* in plasma tissue.** The pQTL weights were plotted in the y-axis for all test genetic variants in the test gene region, with color-coded with respect to -log10 (GWAS p-value). Test SNPs with GWAS p-value <${10}^{-5}$ were colored.

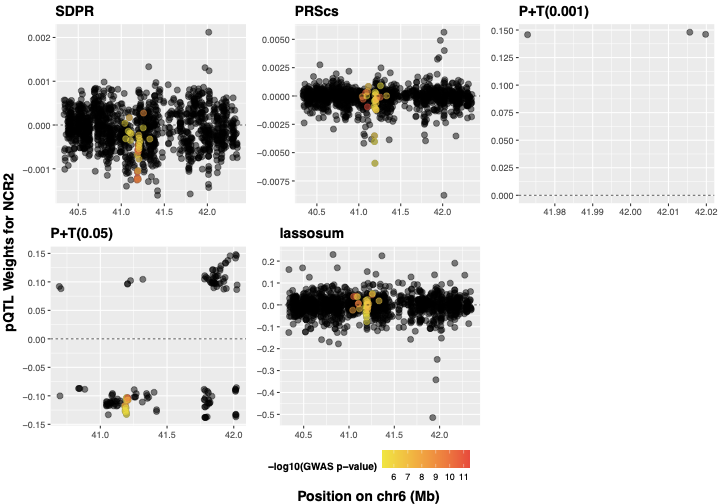

**Fig. S7. Manhattan plots of TWAS results of AD dementia by three Tools: TIGAR/DPR, PrediXcan/Elastic_Net and FUSION/BestModel.**

Reference transcriptomic data profiled from DLPFC tissue of ROS/MAP cohorts were used. The -log10(q-values) were plotted on the y-axis, and -log10(0.05) was plotted as the dashed horizontal line. Independent significant genes are labeled.

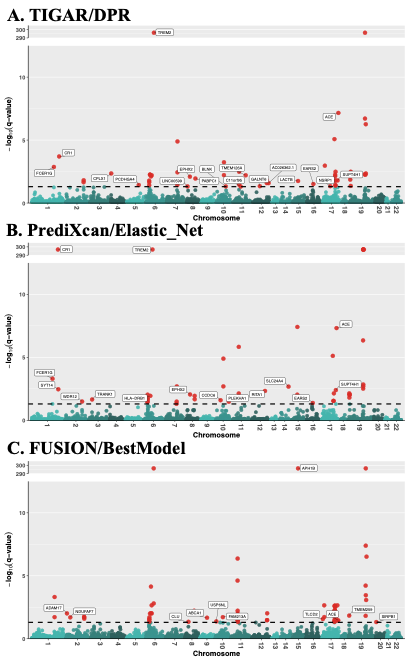
